# Six-Month Dienogest Therapy Reduced the Endometrioma Size, Pelvic Pain, And CA-125 Levels

**DOI:** 10.64898/2026.03.20.26348926

**Authors:** Seyma Ozcan, Ozlem Karabay Akgul, Hakan Guraslan

## Abstract

**Background:** Endometriosis affects approximately 10% of women of reproductive age and is associated with pelvic pain, infertility, and reduced quality of life. Dienogest is widely used for medical management. This study evaluated the effects of dienogest on endometrioma size, serum CA-125 levels, and pelvic pain.

**Methods:** In this retrospective study, medical records of 45 women aged 18–49 years who received oral dienogest (2 mg/day) for at least six months were reviewed. Endometrioma size was assessed by ultrasound, pelvic pain using the Visual Analog Scale (VAS), and serum CA-125 levels from laboratory records. Baseline and six-month values were compared using the Wilcoxon test and correlations were analyzed using Spearman’s test.

**Results:** After six months of treatment, significant reductions were observed in endometrioma size and VAS scores (*p*<0.001) and CA-125 levels (*p*<0.001) compared with baseline. No significant correlation was found between endometrioma size and VAS scores or CA-125 levels either before or after treatment (p>0.05). A significant negative correlation was identified between patient age and post-treatment endometrioma size (r = −0.320, *p*<0.05).

**Conclusion:** Six months of dienogest therapy was associated with significant improvements in lesion size, pain, and biochemical markers. Dienogest may represent an effective medical treatment option for symptomatic patients, particularly for those seeking to avoid surgery and preserve ovarian reserve.

## INTRODUCTION

Endometriosis is a chronic, estrogen-dependent inflammatory disease characterized by the presence of endometrial-like tissue outside the uterine cavity and affects approximately 10% of women of reproductive age (Zondervan et al., 2018; Zondervan et al., 2020). It is a major cause of dysmenorrhea, dyspareunia, chronic pelvic pain, and infertility, significantly impairing quality of life and work productivity (Zondervan et al., 2020; Giudice & Kao, 2004). Ovarian endometriomas are among the most common manifestations of the disease and are frequently associated with pelvic pain, infertility, and reduced ovarian reserve (ESHRE Guideline, 2022; Dunselman et al., 2014).

Management of endometriosis should be individualized based on symptom severity, disease extent, age, and reproductive plans. Although surgery may be effective in selected cases, repeated surgical interventions may lead to adhesions and decreased ovarian reserve (Dunselman et al., 2014; ACOG, 2010). Therefore, long-term medical therapy aimed at symptom control and suppression of disease activity has become a cornerstone of management, particularly in patients who wish to preserve fertility.

Dienogest, a fourth-generation oral progestin, exerts antiproliferative, anti-inflammatory, and anti-angiogenic effects on endometriotic lesions (Foster & Wilde, 1998; McCormack, 2010). Previous studies have demonstrated its effectiveness in reducing endometriosis-related pain and improving patient-reported outcomes (Andres et al., 2015; Römer, 2018; Sağlık Gökmen et al., 2023). In addition to symptomatic improvement, suppression of lesion activity may result in reductions in endometrioma size and serum CA-125 levels, which may reflect disease activity in moderate to severe cases.

Although several studies have reported the clinical benefits of dienogest, real-world data simultaneously evaluating its effects on endometrioma size, pelvic pain severity, and CA-125 levels remain limited. Assessment of these parameters together may provide a more comprehensive evaluation of treatment response in routine clinical practice. Therefore, the aim of this study was to investigate the effects of six months of dienogest therapy on endometrioma size, pelvic pain assessed by the Visual Analog Scale (VAS), and serum CA-125 levels in patients with endometriosis.

## METHODS

### Study design and setting

This retrospective study was conducted in the Department of Obstetrics and Gynecology at a tertiary care training and research hospital in Istanbul, Turkey. Medical records of patients followed in the endometriosis and chronic pelvic pain outpatient clinic were reviewed. Clinical data at presentation were compared with findings obtained after treatment.

### Study population

Patients aged 18–49 years with a diagnosis of endometriosis who had received regular oral dienogest therapy (2 mg/day) for at least six months were eligible for inclusion. A total of 45 patients who met the predefined inclusion and exclusion criteria were included in the final analysis. Patients were excluded if they were younger than 18 or older than 49 years, had known or suspected breast cancer, a history of venous thromboembolism, active smoking status, a history of malignancy, known liver disease, or diabetes mellitus with cardiovascular complications. Clinical and laboratory data were retrospectively obtained from the hospital information system (HIS) and outpatient clinic records.

### Ultrasound assessment

Endometrioma measurements were performed using transvaginal ultrasonography with a 5-MHz probe (Esaote). In virgin patients, transabdominal ultrasonography was used. All measurements were performed by two experienced gynecologists with more than 10 years of experience in the endometriosis unit. The largest diameter (mm) of the dominant endometrioma was recorded and used for analysis.

### Outcome measures

Pelvic pain assessment: Pelvic pain severity was evaluated using the Visual Analog Scale (VAS), ranging from 0 (no pain) to 10 (worst imaginable pain). VAS scores recorded at baseline and after at least six months of treatment were compared.

**Figure 1:**
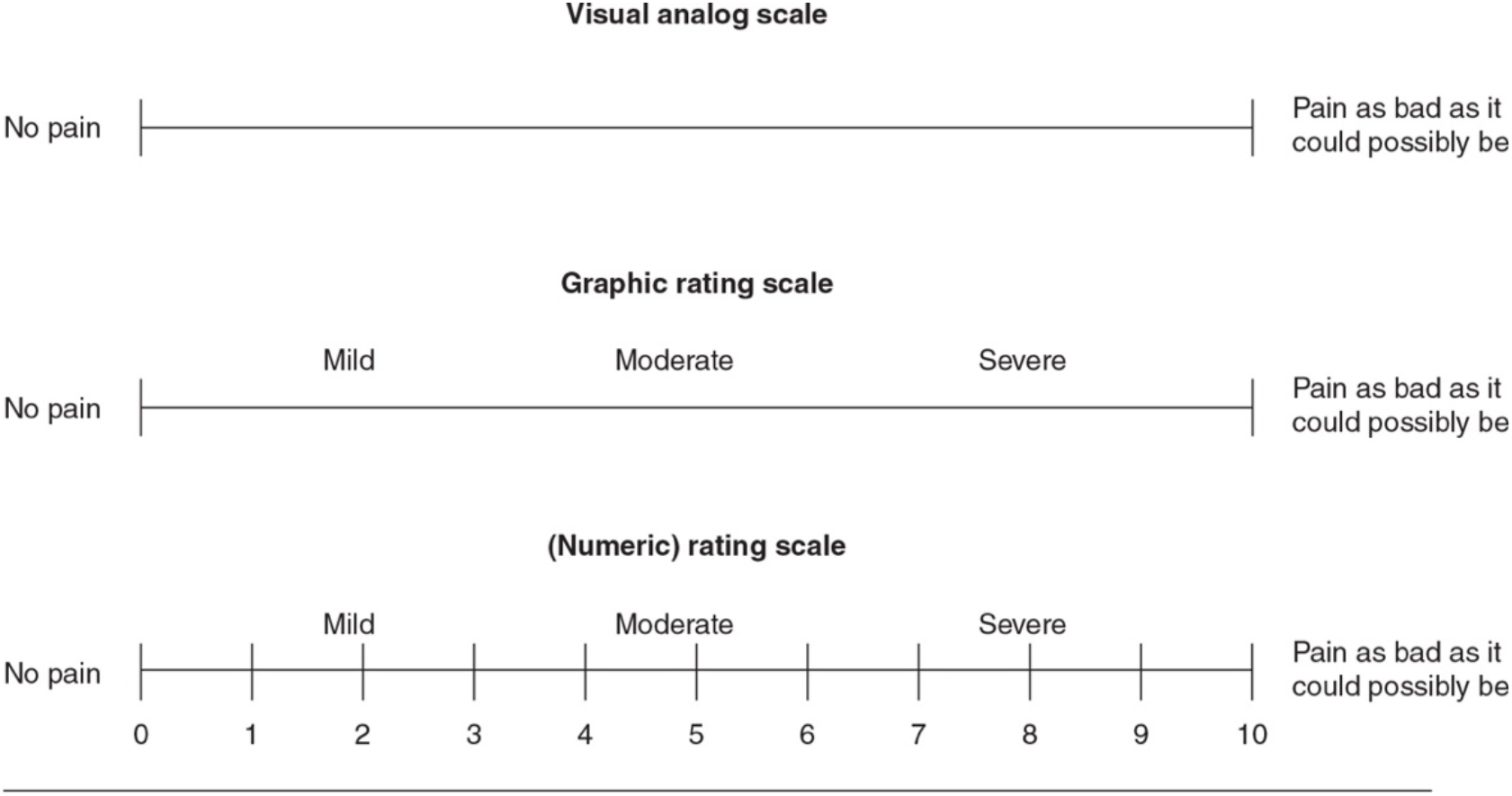
Visual analog scale with numeric pain rating scale. used in this study. Pain intensity was assessed using a 0–10 numeric rating, where 0 indicates no pain, and 10 indicates the worst imaginable pain. Participants were asked to select the number that best represented their perceived pain level at the time of assessment. This figure has been copied from Gift A.G., 1989.

Serum CA-125: Serum CA-125 levels were obtained from hospital laboratory records at baseline and after treatment. Changes in CA-125 levels were used to assess the biochemical response to therapy.

Endometrioma size: Endometrioma size was evaluated using ultrasonography before treatment and after at least six months of therapy. The maximum diameter of the largest endometrioma was recorded in millimeters.

### Statistical analysis

Descriptive statistics for continuous variables were presented as mean ± standard deviation, median, minimum, and maximum values, while categorical variables were expressed as numbers and percentages. Normality of continuous data was assessed using the Shapiro–Wilk test.

Comparisons between baseline and six-month values for VAS score, CA-125 level, and endometrioma size were performed using the Wilcoxon signed-rank test. Z values represent the test statistics of the Wilcoxon analysis. VAS scores range from 0 (no pain) to 10 (worst imaginable pain). Endometrioma size is expressed in millimeters (mm) and CA-125 levels in units per milliliter (U/mL). A p-value < 0.05 was considered statistically significant.

Correlations between endometrioma size, VAS score, and CA-125 levels were analyzed using Spearman’s correlation coefficient. Spearman’s correlation coefficients (r) were calculated to assess the relationship between endometrioma size and VAS scores and serum CA-125 levels at baseline and post-treatment. A p-value < 0.05 was considered statistically. In addition, as indicated above, Spearman’s rank correlation analysis was performed to assess the association between patient age and clinical parameters at baseline and after six months of treatment. Correlation coefficients (r) and corresponding p-values are presented. A p-value < 0.05 was considered statistically significant.

Statistical analyses were performed using IBM SPSS for Windows version 20.0 (SPSS Inc., Chicago, IL, USA). A p-value <0.05 was considered statistically significant.

## RESULTS

In this study, 45 patients with endometrioma were evaluated. The mean age of the patients was 35 years (range: 23–47 years). Chronic comorbidities were present in 4.4% of patients (n = 2). Among these, one patient had asthma, and the other had hyperprolactinemia. Medication use was reported in 4.4% of patients (n = 2); one patient was using an inhaler, and the other was receiving cabergoline (Dostinex). No personal or family history of malignancy was identified in the study population. Baseline demographic and clinical characteristics are presented in Table 1, where age is expressed as mean ± standard deviation and median (range), and chronic disease and medication use are reported as numbers (n) and percentages (%) (Table 1).

**Table 1.**
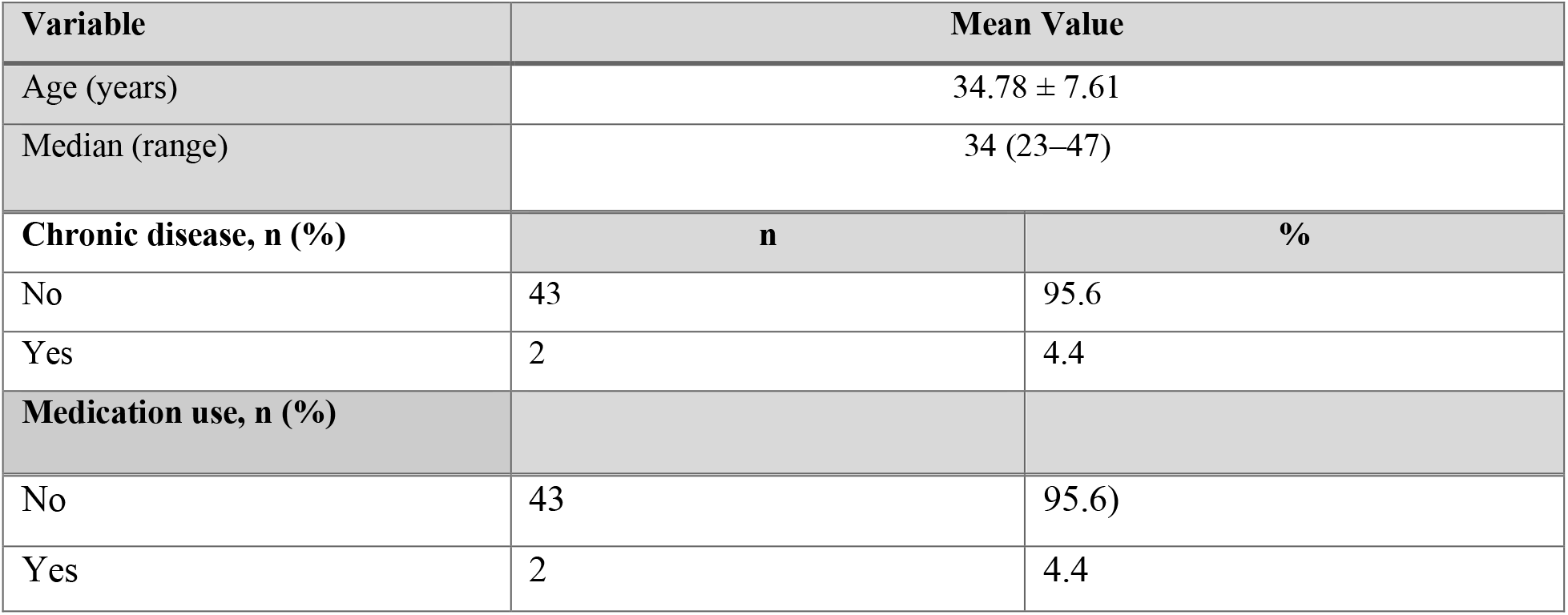
Baseline characteristics of the study population (n = 45).

We observed a significant reduction in pelvic pain, serum CA-125 levels, and endometrioma size after six months of dienogest therapy (Table 2). After six months of dienogest therapy, significant decreases were observed in all evaluated clinical and radiological parameters (Table 2). A significant difference was observed between baseline and six-month VAS scores (p < 0.001). VAS scores at six months were significantly lower compared to pretreatment values (Table 2). Serum CA-125 levels were also significantly reduced following six months of dienogest treatment. A significant difference was observed between baseline and six-month CA-125 levels (p < 0.01) (Table 2). Similarly, endometrioma size showed substantial reduction after treatment. The median lesion size decreased from 45 mm to 35 mm. Endometrioma size at six months was significantly smaller compared to pretreatment measurements (Table 2).

**Table 2.** Comparison of VAS scores, CA-125 levels, and endometrioma size at baseline and at six months of treatment.

|  | Before treatment | Post Treatment | Test statistics | p |
| --- | --- | --- | --- | --- |
|  | Baseline | 6 Months |  |  |
| | Mean ± SD<br>Median (Min-Max) | Mean ± SD<br>Median (Min-Max) | | |
| VAS | 6.69±2.25<br>7 (0-10) | 3.64±2.55<br>3 (0-10) | Z=-4.881 | <0.001<sup>d</sup> |
| CA-125 (U/mL:) | 61.85±58.25<br>41 (8-363) | 39.02±29.95<br>31 (8-146) | Z=-3.058 | 0.002<sup>d</sup> |
| Endometrioma size (mm) | 50.22±22.77<br>45 (18-118) | 38.24±20.43<br>35 (16-118) | Z=-4.216 | <0.001<sup>b</sup> |

*d: Wilcoxon test*

We then evaluated the relationship between endometrioma size and clinical parameters to determine whether lesion size was associated with pain severity or serum CA-125 levels. No significant correlation was found between baseline endometrioma size and baseline VAS scores (p > 0.05). Similarly, no significant correlation was observed between baseline endometrioma size and baseline CA-125 levels (p > 0.05) (Table 3). Likewise, no significant correlation was identified between post-treatment endometrioma size and post-treatment VAS scores (p > 0.05). No significant association was observed between post-treatment endometrioma size and post-treatment CA-125 levels (p > 0.05) (Table 3).

**Table 3.** Correlation between endometrioma size and clinical parameters at baseline and after six months of treatment.

|  | Endometrioma size before treatment |  |
| --- | --- | --- |
|  | **r\*** | **p** |
| **Pre-treatment VAS** | 0.010 | 0.949 |
| **Post-treatment CA-125** | 0.152 | 0.319 |
|  | Endometrioma size after treatment |  |
|  | **r\*** | **p** |
| **Pre-treatment VAS** | 0.283 | 0.073 |
| **Post-treatment CA-125** | 0.216 | 0.175 |

*\*Spearman's Correlation Coefficient*

**Table 4:** Correlation between patient age and baseline and post-treatment endometrioma size, VAS scores, and CA-125 levels.

|  | Age |  |
| --- | --- | --- |
| | $r^*$ | p |
| Baseline VAS | -0.072 | 0.638 |
| Baseline CA-125 | 0.008 | 0.959 |
| Baseline Endometrioma Size | -0.143 | 0.347 |
| Post-treatment VAS | -0.147 | 0.336 |
| Post-treatment CA-125 | -0.249 | 0.099 |
| Post-treatment Endometrioma Size | -0.320 | 0.041 |

*\*Spearman's Correlation Coefficient*

Afterwards, we examined whether patient age was associated with baseline and post-treatment clinical parameters. No significant correlation was found between patient age and baseline endometrioma size, VAS scores, or CA-125 levels (p>0.05). Similarly, no significant correlation was observed between patient age and post-treatment VAS scores or CA-125 levels (p>0.05). However, a significant negative correlation was identified between patient age and post-treatment endometrioma size (r = −0.320, p<0.05), indicating that endometrioma size decreased with increasing age.

## DISCUSSION

In this study, six months of treatment with 2 mg/day oral dienogest in 45 patients with endometrioma resulted in significant reductions in endometrioma size, pelvic pain severity (VAS score), and serum CA-125 levels. However, no correlation was observed between endometrioma size and VAS score or CA-125 levels. These findings suggest that while dienogest effectively reduces disease activity and symptom burden, lesion size alone may not directly reflect symptom severity or biochemical markers.

Symptom control remains the primary goal in the management of endometriosis (Ferrero et al., 2018). Current guidelines from the American Society for Reproductive Medicine recommend medical therapy as the first-line approach for patients with superficial or persistent disease, reserving surgical treatment for those with large endometriomas or disease refractory to medical management (ASRM, 2014). Effective medical therapies that reduce symptoms and suppress lesion activity are therefore essential, particularly for patients who wish to avoid surgery and preserve ovarian reserve.

Previous studies have not clearly demonstrated superiority between estrogen–progestin combinations and progestin-only therapies in terms of efficacy, safety, tolerability, or cost (Vercellini et al., 2016). Some patients using combined oral contraceptives may experience pain during hormone-free intervals due to withdrawal bleeding; in such cases, continuous rather than cyclic use is recommended (Zorbas et al., 2015; Vercellini et al., 2018). Because endometriosis is a chronic disease requiring long-term treatment, progestins may be preferred as first-line therapy in patients who cannot tolerate or have contraindications to combined oral contraceptives (Vercellini et al., 2016).

Dienogest is a selective progestin that suppresses local estrogen production and inhibits the proliferation of endometrial cells. It also partially suppresses hypothalamic GnRH secretion, leading to decreased LH and FSH levels and reduced ovarian estrogen production. These mechanisms create a hypoestrogenic environment that limits the growth and activity of ectopic endometrial tissue. Dienogest has no androgenic, glucocorticoid, or mineralocorticoid activity and is characterized by high oral bioavailability and a pharmacokinetic profile suitable for once-daily administration (McCormack et al., 2010).

The effectiveness of dienogest in reducing endometriosis-associated pain has been demonstrated in several clinical studies. Römer et al. (2018) reported significant long-term reductions in pelvic pain scores in women receiving 2 mg/day dienogest, both after surgery and as primary treatment. Similarly, Sağlık Gökmen et al. (2023) evaluated 64 patients treated with dienogest and reported significant reductions in both endometrioma size and pain scores. In our study, mean endometrioma size decreased from 50.2 mm to 38.2 mm, and the mean VAS score decreased from 6.69 to 3.64, supporting the lesion-suppressive and symptom-relieving effects of therapy.

Chen et al. (2024) also demonstrated significant reductions in pain scores, endometrioma size, and CA-125 levels after six months of dienogest treatment. In their study, endometrioma size decreased by approximately 35% after six months, and CA-125 levels showed marked improvement. Our findings of reduced CA-125 levels and decreased lesion size are consistent with these observations. These effects may be attributed to dienogest-induced stromal atrophy within endometriotic lesions, decreased cellular proliferation, and a consequent reduction in active ectopic endometrial tissue. Furthermore, as endometriosis is associated with peritoneal irritation and inflammation, the anti-inflammatory effects of dienogest may contribute to the observed decline in serum CA-125 levels.

The strength of the present study is the simultaneous evaluation of clinical, radiological, and biochemical treatment outcomes in a real-world clinical setting. Assessing endometrioma size, pain severity, and CA-125 levels together provides a more comprehensive assessment of treatment response.

However, several limitations should be acknowledged. The retrospective design, relatively small sample size, and absence of a control group limit the generalizability of the findings. In addition, the follow-up period was limited to six months, and long-term outcomes were not evaluated. Larger prospective studies with longer follow-up are needed to confirm these results.

Overall, our findings suggest that dienogest is an effective medical treatment option for patients with endometriosis, providing significant improvements in symptoms and disease-related parameters while potentially reducing the need for surgical intervention.

## CONCLUSION

In this study, six months of dienogest therapy was associated with significant reductions in endometrioma size, pelvic pain severity, and serum CA-125 levels. These findings support the use of dienogest as an effective and well-tolerated treatment option that may delay the need for surgical intervention, particularly in patients who wish to preserve fertility.

## LIMITATION OF STUDY

The relatively small sample size, retrospective design, and short follow-up period limit the generalizability of these findings. Future prospective, randomized studies with larger patient populations and longer follow-up are needed to confirm the long-term efficacy and safety of dienogest therapy.

## Data Availability

All data produced in the present work are contained in the manuscript.

## ACKNOWLEDGMENTS

We would like to thank the staff of the Endometriosis and Chronic Pelvic Pain Clinic at Bagcılar Training and Research Hospital for their support in patient follow-up and data management.

## ETHICS/IRB STATEMENT

The study was approved by the Clinical Research Ethics Committee of the University of Health Sciences Bagcılar Training and Research Hospital. Due to the retrospective design, informed consent was waived.

## DATA AVAILABILITY STATEMENT

Data are available from the corresponding author upon reasonable request.

